# OVM-200, a recombinant overlapping peptide vaccine, breaks immune tolerance to survivin in patients with advanced solid tumours: a multicentre, open-label, dose-expansion, phase 1 study

**DOI:** 10.64898/2026.11.08.26367556

**Authors:** Peter Kok-Ting Wan, Thomas Morris, Martin D. Forster, Tianen Zhang, Michael Flynn, Florence Chamberlain, Mark Tuthill, Fiona Thistlethwaite, Anja Williams, William Finch, Wenshu Lu, Shisong Jiang

## Abstract

Survivin (BIRC5) is broadly overexpressed in human cancers but, as a self-antigen, is subject to immune tolerance that limits spontaneous antitumour immunity. OVM-200 is a recombinant overlapping peptide vaccine spanning full-length survivin, with cathepsin S-cleavable linker sequences inserted between adjacent peptides to enhance antigen processing and presentation. OVM-200-100 was a multicentre, open-label phase 1 study conducted at 4 UK sites. The pooled 2,000-μg cohort comprised 27 patients with advanced non-small cell lung, ovarian or prostate cancer, including 24 patients from phase 1b and 3 treated at this dose in phase 1a. OVM-200 was administered subcutaneously every 2 weeks for 3 initial vaccinations, with up to 8 additional doses in patients deriving clinical benefit. The primary endpoint was safety and tolerability, secondary endpoints included survivin-specific immunogenicity, and antitumour activity by RECIST v1.1 was exploratory. OVM-200-related adverse events were exclusively grade 1-2, with no treatment-related grade ≥3 adverse events, dose-limiting toxicities, serious adverse events or deaths. A systematic literature analysis showed low spontaneous anti-survivin seroreactivity in patients with cancer. In our cohort, pretreatment tumours expressed survivin despite low baseline survivin-specific immunity. OVM-200 vaccination subsequently induced robust humoral and cellular responses, with median anti-survivin IgG titres increasing from 20 at baseline to a peak of 81,920 and median interferon-γ ELISpot responses increasing from 16.7 to 504.6 spot-forming units per 10⁶ PBMCs. Patients receiving ≥4 vaccinations developed higher anti-survivin antibody titres and stronger T cell responses than those receiving fewer doses. Among 24 RECIST-evaluable patients, the best overall response was stable disease (SD) in 6 patients (25%). Patients with SD had significantly higher anti-survivin antibody titres than those with progressive disease (PD) (p=0.0418). OVM-200 was well tolerated and induced robust survivin-specific immunity, providing clinical evidence that it can break immune tolerance to self-antigen survivin and supporting further development. ClinicalTrials.gov identifier: NCT05104515.

## Introduction

Therapeutic cancer vaccines aim to induce tumour-specific immune responses capable of controlling or eliminating established malignancies^1^. Recent clinical studies have demonstrated encouraging activity for personalised neoantigen vaccines, highlighting the potential of vaccination as an effective cancer immunotherapy^2–5^. However, personalised vaccines require tumour sequencing and individualised manufacturing, limiting their scalability, cost-effectiveness, and timely delivery. Consequently, there remains a substantial need for effective off-the-shelf cancer vaccines. Most off-the-shelf cancer vaccines target tumour-associated antigens (TAAs), which are shared across patients and tumour types^6^. Unlike tumour-specific antigens (TSAs), however, TAAs are self-antigens and are therefore subject to central and peripheral immune tolerance, resulting in weak endogenous immune responses and limited clinical efficacy^7,8^. Overcoming immune tolerance remains a major challenge in the development of TAA-based vaccines.

Survivin (baculoviral IAP repeat-containing protein 5 [BIRC5]) is a tumour-associated antigen (TAA) and multifunctional oncofetal protein that is overexpressed in a broad range of human malignancies but shows limited expression in most differentiated adult tissues. It plays key roles in cell division, inhibition of apoptosis, and tumour progression, and its overexpression has been associated with treatment resistance and poor clinical outcomes^9–11^. These characteristics make survivin an attractive target for therapeutic cancer vaccination^12,13^. However, because survivin is a non-mutated self-antigen, endogenous immune responses against it are constrained by central and peripheral tolerance, which has limited the clinical efficacy of conventional survivin-targeted vaccines^14^.

Previous survivin-based vaccines have demonstrated that survivin can be safely targeted and that survivin-specific immune responses can be induced in patients with cancer^15–17^. In particular, the survivin peptide-based vaccine SurVaxM has demonstrated favourable safety, immunogenicity and encouraging clinical activity in patients with recurrent malignant glioma^16^. However, like many peptide-based survivin vaccines, SurVaxM targets a limited number of predefined epitopes and relies on presentation by selected HLA haplotypes, potentially restricting breadth of immune responses and limiting applicability across genetically diverse patient populations^16,18,19^. Vaccine platforms capable of presenting the full repertoire of survivin epitopes in an HLA-unrestricted manner may therefore broaden immune responses and extend the potential clinical benefit of survivin-targeted immunotherapy.

To overcome the limitations of conventional TAA-based and epitope-based peptide vaccines, we developed the recombinant overlapping peptide (ROP) antigen engineering platform. Rather than selecting a limited number of predefined epitopes, the ROP platform re-engineers the full-length antigen, including survivin, into a series of overlapping peptides linked by cathepsin S-cleavable sequences, preserving the complete antigenic repertoire while optimising intracellular antigen processing and presentation^20^. This design is intended to enhance the immunogenicity of self-antigens, broaden CD4⁺ and CD8⁺ T cell recognition in an HLA-unrestricted manner^21^. The disruption of the native three-dimensional structure abolishes the biological activity of the survivin oncoprotein while retaining its antigenicity, potentially improving the safety profile compared with administration of the native oncoprotein.

OVM-200 is the first therapeutic vaccine developed using the ROP antigen engineering platform and the first ROP vaccine to be administered to humans. In the phase 1a dose-escalation study, OVM-200 was administered subcutaneously at doses of 250, 500, 1000, and 2000 μg to patients with advanced solid cancers^22^. OVM-200 demonstrated a favourable safety profile across dose levels, with the 2000 μg dose inducing the strongest survivin-specific humoral and cellular immune responses. Based on these findings, the 2000 μg dose was selected for further evaluation in the phase 1b expansion cohort.

Here, we report the results the pooled 2000 µg cohort of OVM-200-100 study, comprising patients treated at 2000 µg in phase 1a dose-escalation and the phase 1b expansion cohort, in patients with advanced non-small cell lung cancer (NSCLC), ovarian cancer, and prostate cancer. The objectives were to evaluate the safety and tolerability of OVM-200 at the selected dose, to characterise survivin-specific humoral and cellular immune responses, explore the potential association between vaccine-induced immunity and clinical outcome, and assess preliminary antitumour activity.

## Results

### Patients characteristics

Between 15 Oct 2021 and 3 Dec 2025, 29 patients were screened for the phase 1b expansion cohort, of whom 5 patients failed screening, and 24 (83%) were assigned to receive OVM-200 at 2000 µg (Figure 1). Three patients who had received OVM-200 at 2000 µg during phase 1a were included in the pooled 2000 µg analysis population, resulting in a total of 27 patients evaluated. Of the 24 patients enrolled in phase 1b, 11 (46%) completed the trial and 13 (54%) discontinued: 4 died, 1 was lost to follow-up, 2 discontinued due to progressive disease, 2 withdrew consent, and 4 discontinued for other reasons.

**Figure 1.**
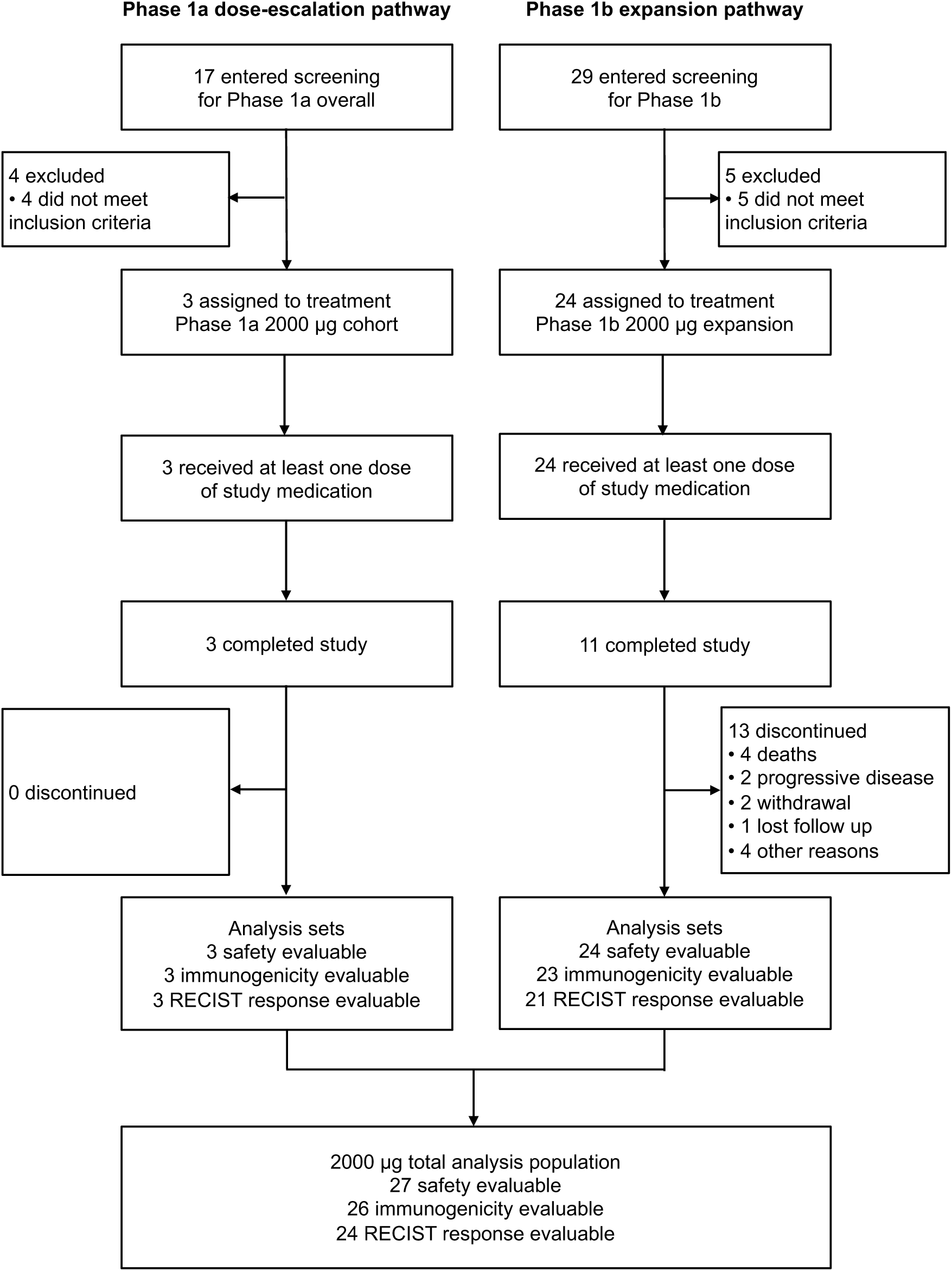
Trial profile. Consort flow diagram for patients receiving OVM-200 at 2000 µg. Patients treated with OVM-200 2,000 µg in phase 1a (n=3) and phase 1b (n=24) are shown separately before merging into the pooled 2,000 µg analysis population (n=27). Analysis populations are shown at the end of each pathway. RECIST, Response Evaluation Criteria in Solid Tumours.

Across the pooled 2000 µg cohort (n=27), 4 patients had NSCLC, 11 had ovarian cancer, and 12 had prostate cancer. At screening, 26 patients (96%) had stage IV disease and 1 (4%) had stage III disease (Table 1). The median age was 63.0 years (range 54-75), and 15 patients (56%) were female. The most common sites of metastasis were bone [13 patients (48%)], lymph nodes [12 (44%)], liver [10 (37%)], lung [6 (22%)], and peritoneum [5 (19%)].

**Table 1.**
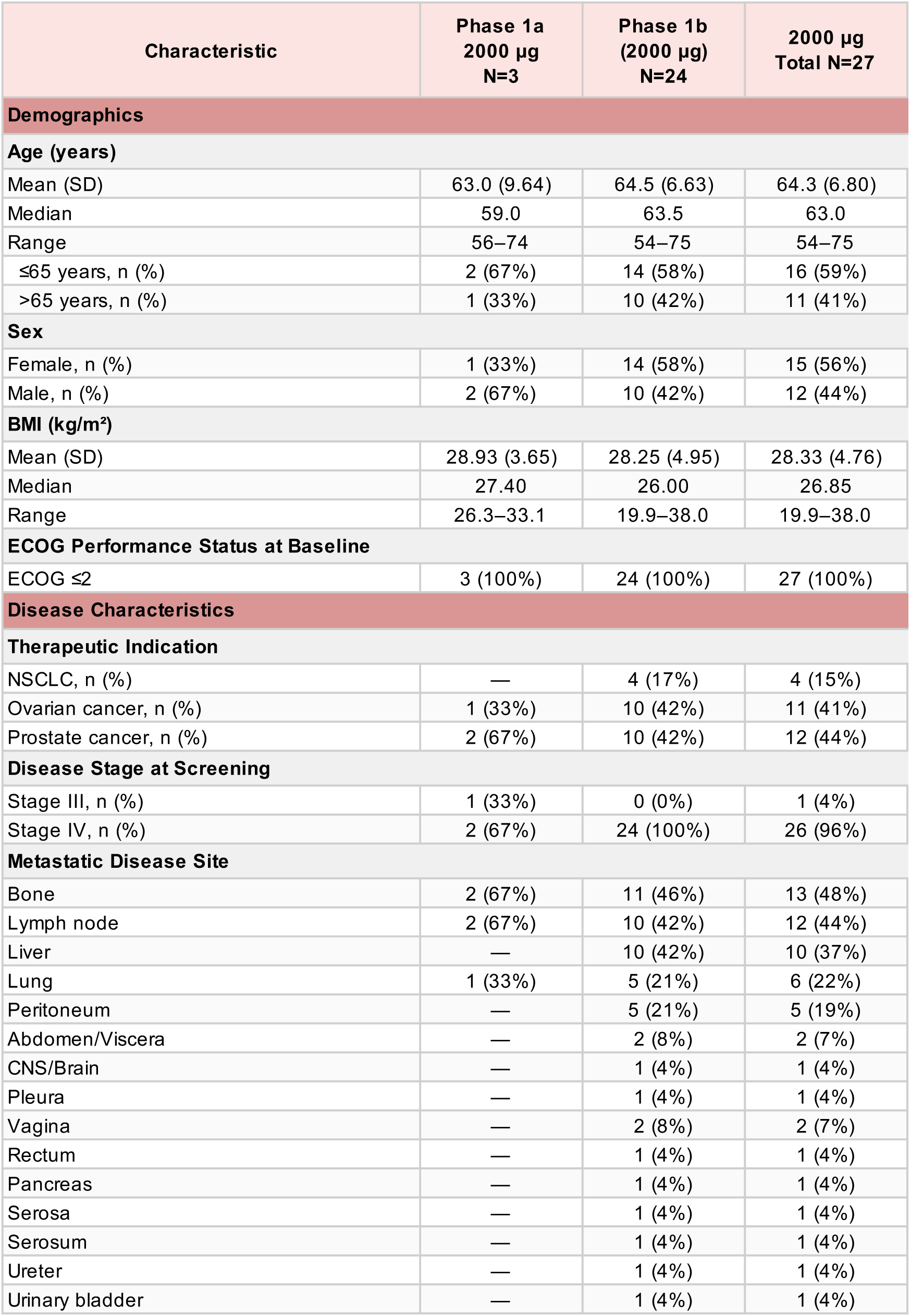
Baseline demographics and disease characteristics. Data are n (%) unless stated. The combined 2000 µg cohort comprised phase 1a 2000 µg (n=3) and phase 1b (n=24) patients. ECOG, Eastern Cooperative Oncology Group performance status; BMI, body mass index; NSCLC, non-small cell lung cancer.

| Characteristic | Phase 1a<br>2000 µg<br>N=3 | Phase 1b<br>(2000 µg)<br>N=24 | 2000 µg<br>Total N=27 |
|---|---|---|---|
| **Demographics** |  |  |  |
| **Age (years)** |  |  |  |
| Mean (SD) | 63.0 (9.64) | 64.5 (6.63) | 64.3 (6.80) |
| Median | 59.0 | 63.5 | 63.0 |
| Range | 56–74 | 54–75 | 54–75 |
| ≤65 years, n (%) | 2 (67%) | 14 (58%) | 16 (59%) |
| >65 years, n (%) | 1 (33%) | 10 (42%) | 11 (41%) |
| **Sex** |  |  |  |
| Female, n (%) | 1 (33%) | 14 (58%) | 15 (56%) |
| Male, n (%) | 2 (67%) | 10 (42%) | 12 (44%) |
| **BMI (kg/m<sup>2</sup>)** |  |  |  |
| Mean (SD) | 28.93 (3.65) | 28.25 (4.95) | 28.33 (4.76) |
| Median | 27.40 | 26.00 | 26.85 |
| Range | 26.3–33.1 | 19.9–38.0 | 19.9–38.0 |
| **ECOG Performance Status at Baseline** |  |  |  |
| ECOG ≤2 | 3 (100%) | 24 (100%) | 27 (100%) |
| **Disease Characteristics** |  |  |  |
| **Therapeutic Indication** |  |  |  |
| NSCLC, n (%) | — | 4 (17%) | 4 (15%) |
| Ovarian cancer, n (%) | 1 (33%) | 10 (42%) | 11 (41%) |
| Prostate cancer, n (%) | 2 (67%) | 10 (42%) | 12 (44%) |
| **Disease Stage at Screening** |  |  |  |
| Stage III, n (%) | 1 (33%) | 0 (0%) | 1 (4%) |
| Stage IV, n (%) | 2 (67%) | 24 (100%) | 26 (96%) |
| **Metastatic Disease Site** |  |  |  |
| Bone | 2 (67%) | 11 (46%) | 13 (48%) |
| Lymph node | 2 (67%) | 10 (42%) | 12 (44%) |
| Liver | — | 10 (42%) | 10 (37%) |
| Lung | 1 (33%) | 5 (21%) | 6 (22%) |
| Peritoneum | — | 5 (21%) | 5 (19%) |
| Abdomen/Viscera | — | 2 (8%) | 2 (7%) |
| CNS/Brain | — | 1 (4%) | 1 (4%) |
| Pleura | — | 1 (4%) | 1 (4%) |
| Vagina | — | 2 (8%) | 2 (7%) |
| Rectum | — | 1 (4%) | 1 (4%) |
| Pancreas | — | 1 (4%) | 1 (4%) |
| Serosa | — | 1 (4%) | 1 (4%) |
| Serosum | — | 1 (4%) | 1 (4%) |
| Ureter | — | 1 (4%) | 1 (4%) |
| Urinary bladder | — | 1 (4%) | 1 (4%) |

For the immunogenicity analysis, 26 patients were evaluable. 1 patient was excluded because evaluable samples were unavailable. The numbers of patients contributing data declined at later timepoints: at week 8, antibody and T cell data were available for 23 and 22 patients, respectively; the corresponding numbers were 14 and 9 at week 16, and 9 and 8 at week 24. For the assessment of antitumour activity, best overall response was available for 24 patients. Radiological assessments were available for 18 patients at week 8, 6 at week 16, and 8 at week 24.

### Safety

All 27 patients in the 2000 µg cohort were included in the safety analysis. OVM-200-related adverse events occurred in all 27 patients (100%) and were grade 1-2 in all cases (Table 2). No OVM-200-related grade ≥ 3 adverse events, dose-limiting toxicities, serious adverse events, or deaths were reported. One patient (1/27, 4%) discontinued OVM-200 because of an OVM-200-related injection-site reaction.

**Table 2.** OVM-200-related treatment-emergent adverse events. Data are n (%). OVM-200-related treatment-emergent adverse events are shown. Grading per CTCAE version 5.0. DLT, dose-limiting toxicity.

| **OVM-200-Related Adverse Event** | **Grade 1-2** | **Grade 3-4** |
|---|---|---|
| **Any OVM-200-related TEAE** |  |  |
| Grade 1 | 27 (100%) | - |
| Grade 2 | 3 (11%) | - |
| Grade 3-4 | - | 0 |
| Leading to discontinuation of OVM-200 | 1 (4%) | 0 |
| Dose-limiting toxicities (DLTs) | 0 | 0 |
| **General disorders & administration site conditions** |  |  |
| Injection site reaction | 17 (63%) | 0 |
| Injection site erythema | 6 (22%) | 0 |
| Fatigue | 3 (11%) | 0 |
| Injection site pruritus | 3 (11%) | 0 |
| Injection site pain | 2 (7%) | 0 |
| Injection site bruising | 1 (4%) | 0 |
| Injection site rash | 1 (4%) | 0 |
| **Gastrointestinal disorders** |  |  |
| Vomiting | 2 (7%) | 0 |
| **Musculoskeletal & connective tissue disorders** |  |  |
| Myalgia | 3 (11%) | 0 |
| **Nervous system disorders** |  |  |
| Headache | 2 (7%) | 0 |

**Table 3.** Antitumour activity of OVM-200 by RECIST version 1.1. Data are n (%) unless stated. RECIST categories are reported at week 8, 16, and 24. Objective response rate (ORR) is presented with 80% and 95% confidence intervals. SD, stable disease; PD, progressive disease; NE, not evaluable; W8, week 8 (day 57); W16, week 16 (day 113); W24, week 24 (day 169).

|  |  | **2000 µg Total**<br>**N = 24** |
|---|---|---|
| **W8** | **n** | **18** |
|  | SD, n (%) | 6 (25.0) |
|  | PD, n (%) | 12 (50.0) |
| **W16** | **n** | **6** |
|  | SD, n (%) | 2 (8.3) |
|  | PD, n (%) | 4 (16.7) |
| **W24** | **n** | **8** |
|  | SD, n (%) | 3 (12.5) |
|  | PD, n (%) | 5 (20.8) |
| **W8, 16, 24** | **ORR (%)** | **0** |
|  | 80% CI for ORR | 0 - 9.15 |
|  | 95% CI for ORR | 0 - 14.2 |
| **Best overall response** | **n** | **24** |
|  | SD, n (%) | 6 (25.0) |
|  | PD, n (%) | 17 (70.8) |
|  | NE, n (%) | 1 (4.2) |

The most common OVM-200-related adverse events were injection-site reaction (17/27, 63%), injection-site erythema (6/27, 22%), fatigue (3/27, 11%), injection-site pruritus (3/27, 11%), and myalgia (3/27, 11%) (Table 2). Other OVM-200-related adverse events included injection-site pain (2/27, 7%), vomiting (2/27, 7%), and headache (2/27, 7%).

Grade 3 TEAEs occurred in 6 patients (22%), and none were considered related to OVM-200 (Supplementary Table 1). These events included vascular device infection, abdominal pain, dehydration, pneumonia, hypotension, gamma-glutamyltransferase increased, peritonitis, anaemia, nausea, vomiting, hypoalbuminaemia, ascites, and pelvic pain. Serious TEAEs occurred in 5 of 27 patients (19%), corresponding to 9 events, none of which were considered related to OVM-200. Overall, the safety profile was dominated by low-grade injection-site reactions and systemic symptoms, with no treatment-related serious toxicity observed.

### Humoral and cellular immunogenicity

Among patients in the 2000 µg cohort, 26 were evaluable for baseline antibody analysis. The median baseline anti-survivin IgG titre was 20 (IQR 10-160) (Figure 2A). Following vaccination, median titres increased to 20,480 (IQR 10,240-81,920) at the first post-vaccination assessment (week 5-8), 163,840 (IQR 17,920-327,680) at week 16, and remained 163,840 (IQR 51,200-491,520) at week 24. Anti-survivin antibody titres were higher than baseline at all post-vaccination timepoints in every evaluable patient. Across the cohort, the median peak antibody titre was 81,920 (IQR 17,920-327,680) (Figure 2B).

**Figure 2.**
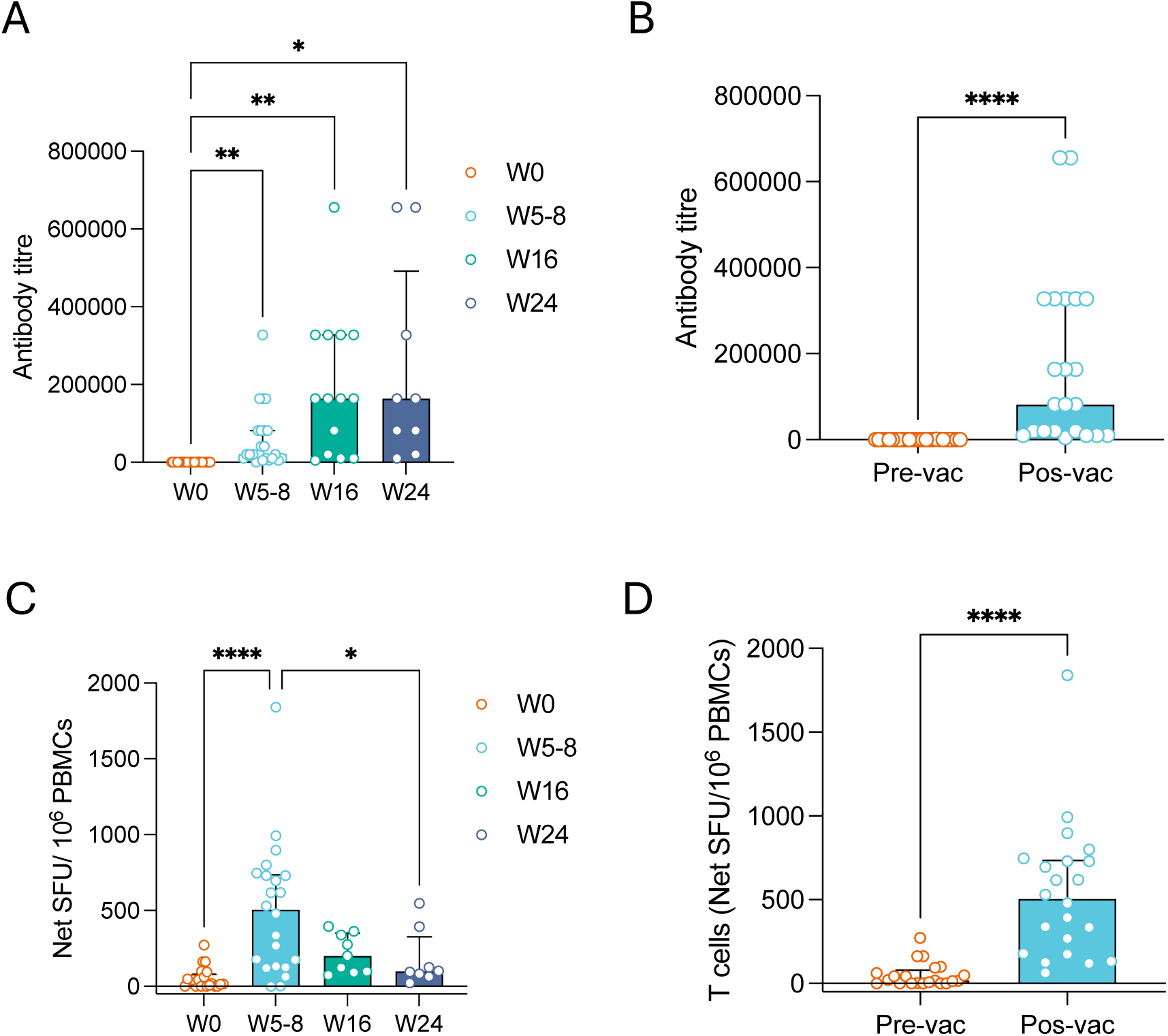
Survivin-specific humoral and cellular immune responses in patients treated with OVM-200. (A) Anti-survivin IgG titres over time in immunogenicity-evaluable patients, shown at baseline (week 0; n=26), early post-vaccination (week 5-8; n=23), week 16 (n=14), and week 24 (n=9). (B) Maximum anti-survivin IgG titre achieved per patient across all timepoints (n=22). (C) Survivin-specific interferon-γ ELISpot responses over time in T cell-evaluable patients, shown at baseline (n=21), early post-vaccination (week 5-8; n=22), week 16 (n=9), and week 24 (n=8). (D) Maximum ELISpot response achieved per patient across all timepoints (n=22). Data are presented as median with interquartile range (IQR).

OVM-200 also induced survivin-specific T cell responses, which peaked at the first post-vaccination assessment with a subsequent decline over time (Figure 2C). 21 patients were evaluable for baseline T cell analysis. The median baseline ELISpot response was 16.7 SFU per 10⁶ PBMCs (IQR 1.0-78.2), increasing to 504.6 SFU per 10⁶ PBMCs (IQR 130.1-734.3) at the first post-vaccination assessment (week 5-8). Median responses subsequently declined to 200.3 SFU per 10⁶ PBMCs (IQR 94.6-350.8) at week 16 and 97.6 SFU per 10⁶ PBMCs (IQR 67.9-326.8) at week 24. In contrast to the durable antibody response, T cell responses showed an early expansion following vaccination, followed by a gradual contraction over time. Across the evaluable cohort, the median peak T cell response was 505.0 SFU per 10⁶ PBMCs (IQR 176.7-734.3) (Figure 2D).

Together, these findings demonstrate that OVM-200 elicited both survivin-specific humoral and cellular immune responses in patients with advanced solid tumours, consistent with its intended mechanism of enhancing immunity against the self-antigen survivin.

### Overcoming pre-existing immune tolerance to survivin

A systematic search was conducted to identify studies evaluating spontaneous survivin-specific immunity in cancer patients. Searches of PubMed (n=677), Web of Science (n=1,286), and Scopus (n=829) identified 2,792 records^23^ (Supplementary Figure 1). Following removal of 1,264 duplicates and five retracted publications, 1,523 records underwent title and abstract screening, of which 1,479 were excluded. 1 study was added during citation searching. Full texts were sought for 45 studies. 4 were unavailable for retrieval, leaving 41 for eligibility assessment. Studies evaluating spontaneous survivin-specific cellular or humoral immunity with solid tumors were initially eligible. At full-text assessment, studies evaluating treatment-induced responses (n=6), or without quantifiable measures of spontaneous survivin-specific immunity (n=1) were excluded. Most cellular immune response studies, and some antibody studies, involved substantial *in vitro* antigen stimulation or lymphocyte expansion and were therefore not considered directly comparable measures of spontaneous *ex vivo* immunity. As only 1 eligible *ex vivo* ELISPOT study remained, the quantitative analysis was restricted to spontaneous survivin-specific humoral immunity measured *ex vivo* by ELISA using titer/dilution-based antibody readouts comparable with the OVM-200 study. 12 studies were included in the final analysis.

Across these 12 studies, comprising 3,001 cancer patients who had not received survivin-directed immunotherapy, spontaneous anti-survivin seroreactivity was generally low^24–35^ (Supplementary Table 2). The median study-level seroprevalence was 15.4% (range 6.0-58.1%), with a patient-weighted pooled prevalence of 14.9% (448/3,001). Prevalence remained low across serum dilutions, with pooled positivity of 11.2% at 1:50 (1 study, n=365), 24.1% at 1:100 (7 studies, n=879) and 11.1% at 1:200 (4 studies, n=1,757).

Spontaneous survivin-specific T cell immunity was also generally low. In metastatic melanoma, only 2/41 (4.9%) patients had detectable baseline survivin-specific responses^36^. In high-risk neuroblastoma, patients (n=9) had a median survivin-specific response of 49 SFU/10⁶ PBMCs^37^. Due to the low number of studies measuring spontaneous survivin-specific T cell responses directly *ex vivo*, a systematic analysis on cellular immunity was not performed.

To investigate whether OVM-200 could induce immune responses against survivin despite pre-existing tumour expression of this self-antigen, a matched exploratory analysis was performed in 12 patients who had both pre-vaccination tumour tissue available for immunohistochemistry and evaluable immunogenicity data. Immunohistochemical analysis demonstrated survivin expression in all 12 tumours before vaccination, including high (+++) expression in 6 patients, moderate (++) expression in 4 patients, and low (+) expression in 2 patients (Figure 3A), confirming widespread expression of the vaccine target antigen. Despite constitutive survivin expression in tumour tissue, baseline survivin-specific humoral immunity was minimal across all expression groups (Figure 3B). Median baseline anti-survivin antibody titres were 15 (IQR 10-160) in the survivin-high group, 20 (IQR 12.5-245) in the survivin-moderate group, and 60 in the survivin-low group.

**Figure 3.**
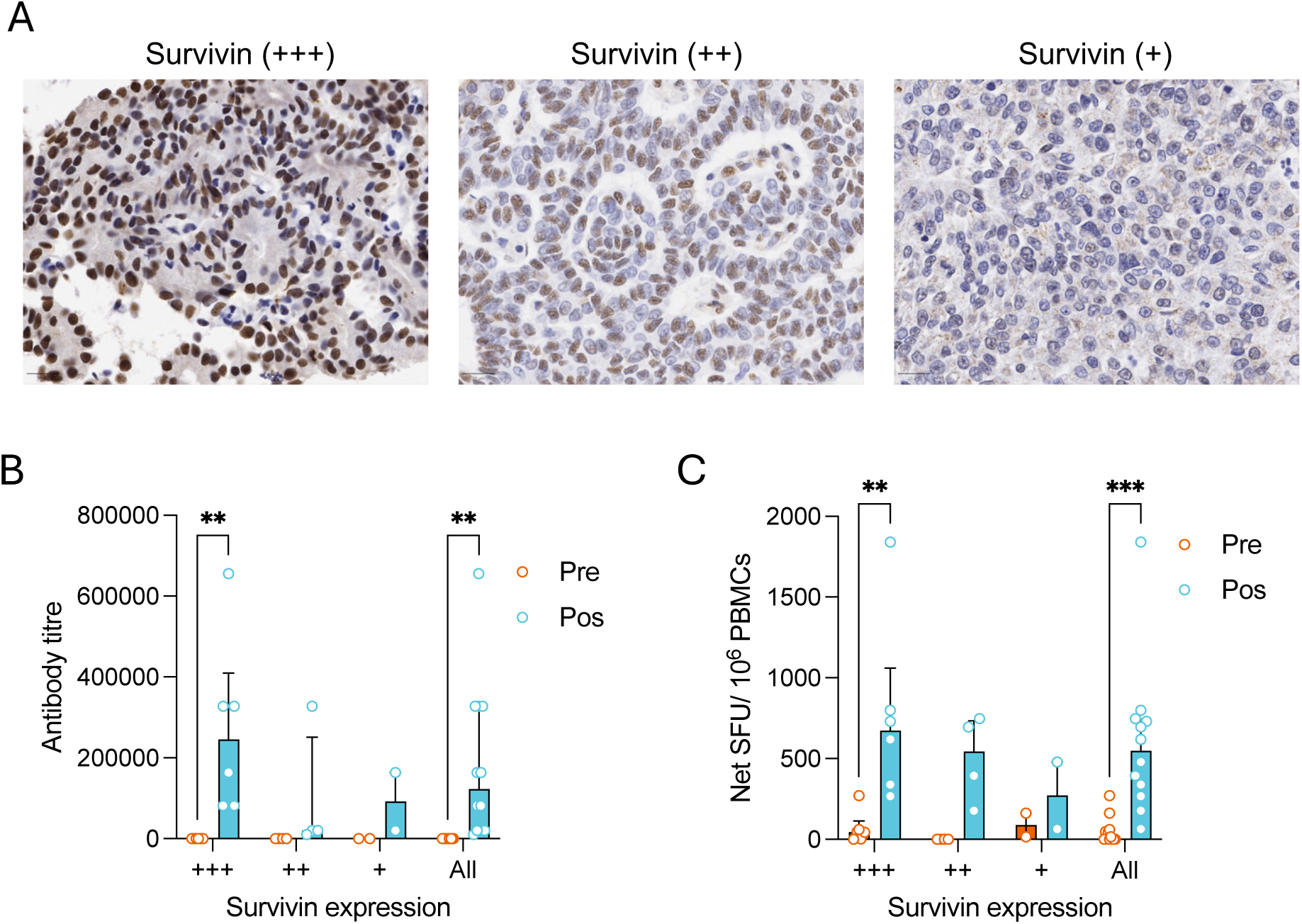
Survivin expression in tumour and the associated anti-survivin immune responses in patients treated with OVM-200. (A) Representative immunohistochemistry images showing tissues classified as having high (+++), moderate (++), or low (+) survivin expression. (B) Anti-survivin IgG titres at pre-vaccination (+++, n=6; ++, n=4; +, n=2) and maximum post-vaccination titre (+++, n=6; ++, n=4; +, n=2), stratified by survivin expression in tumour. (C) Interferon-γ ELISpot responses at pre-vaccination (+++, n=6; ++, n=4; +, n=2) and maximum post-vaccination response (+++, n=6; ++, n=4; +, n=2) by survivin expression. Data are presented as median with interquartile range (IQR). PBMCs, peripheral blood mononuclear cells; SFU, spot-forming units.

Following vaccination with OVM-200, antibody titres increased markedly irrespective of tumour survivin expression, with median peak titres of 245,760 (IQR 81,920-409,600; p=0.0079), 20,480 (IQR 12,800-250,880), and 92,160 in the survivin-high, survivin-moderate and survivin-low expression groups, respectively (Figure 3B).

Similarly, baseline survivin-specific T cell responses were low despite detectable tumour survivin expression, with median pre-vaccination ELISpot responses of 45.8 (IQR 2.2-113.8), 0.4 (IQR 0.0-1.0), and 89.2 SFU per 10⁶ PBMCs in the survivin-high, survivin-moderate and survivin-low expression groups, respectively (Figure 3C). Vaccination induced robust T cell responses across all expression groups, with median peak responses of 674.4 (IQR 320.8-1060.0; p=0.0009), 544.5 (IQR 231.4-733.8), and 272.0 SFU per 10⁶ PBMCs, respectively.

Taken together, these findings demonstrate that, although survivin was constitutively expressed in patients’ tumours before vaccination, spontaneous survivin-specific humoral and cellular immunity remained minimal. Vaccination with OVM-200 elicited robust survivin-specific antibody and T cell responses regardless of tumour survivin expression level, supporting its ability to overcome pre-existing functional immune tolerance to the self-antigen survivin.

### Number of vaccinations and associated immunogenicity

Exploratory analyses suggested that repeated immunisation was associated with stronger vaccine-induced immune responses. Patients were divided into two groups by the number of vaccinations: 1-3 vaccinations (n=11) and ≥4 vaccinations (n=12). Anti-survivin antibody titres increased in both groups, rising from a median of 50 (IQR 10-160) to 15,360 (IQR 10,240-327,680) after 1-3 vaccinations (n=11), and from 25 (IQR 10-160) to 163,840 (IQR 81,920-327,680) after ≥4 vaccinations (n=12). Patients receiving ≥4vaccinations achieved markedly higher peak antibody titres than those receiving only 1-3 vaccinations (Figure 4A).

**Figure 4.**
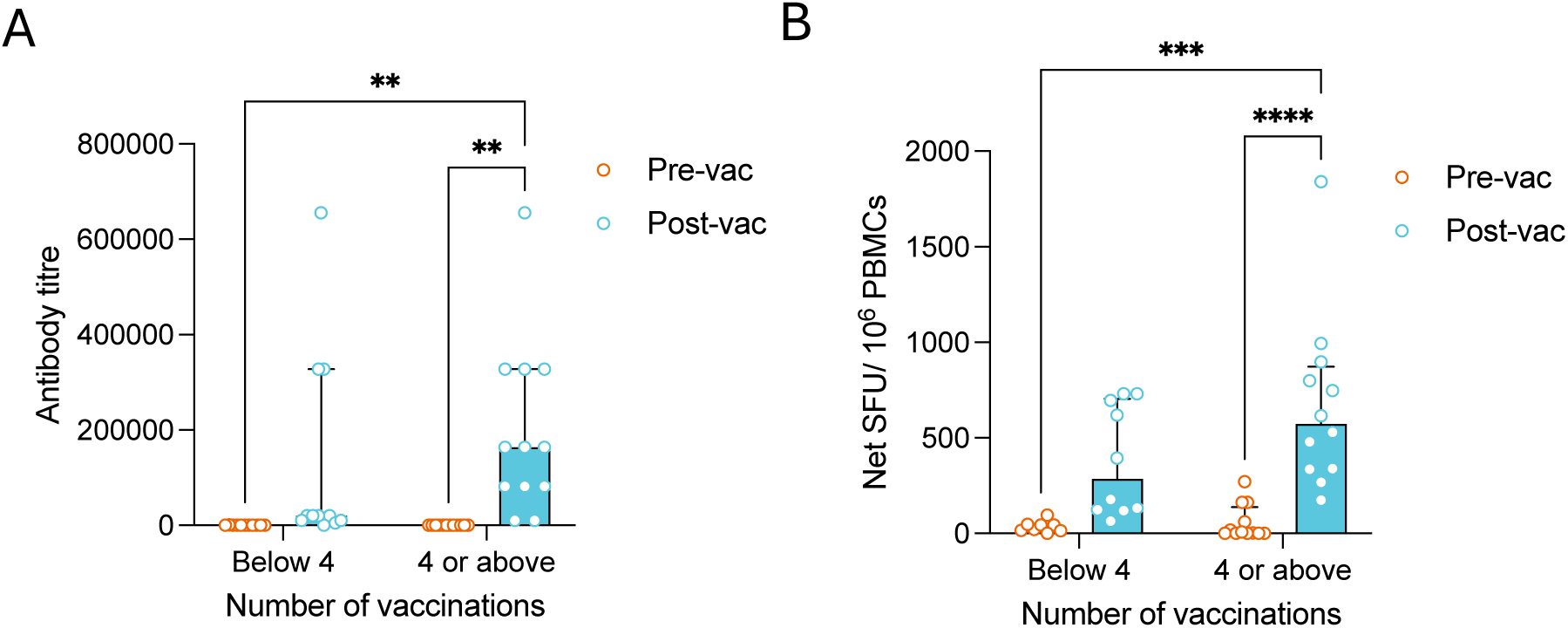
Effect of number of OVM-200 vaccinations on immune responses. Anti-survivin immune responses stratified by number of OVM-200 vaccinations received: 1-3 vaccinations and 4 or more vaccinations. (A) Anti-survivin IgG titres before (1-3 vaccinations, n=11; ≥4 vaccinations, n=12) and after vaccination (1-3 vaccinations, n=11; ≥4 vaccinations, n=12) by vaccination group. (B) Interferon-γ ELISpot responses before (1-3 vaccinations, n=8; ≥4 vaccinations, n=12) and after vaccination (1-3 vaccinations, n=10; ≥4 vaccinations, n=12) by vaccination group. Data are presented as median with interquartile range (IQR). PBMCs, peripheral blood mononuclear cells; SFU, spot-forming units.

Similarly, survivin-specific T cell responses increased following vaccination in both groups. Median ELISpot responses increased from 18 (IQR 13.9-47.7) to 286 (IQR 123.2-703.8) SFU per 10⁶ PBMCs after 1-3 vaccinations (n=10), and from 7 (IQR 1.0-162.6) to 573.0 (IQR 335.8-873.1) SFU per 10⁶ PBMCs after ≥4 vaccinations (n=12) (Figure 4B).Taken together, these exploratory analysis suggest that additional booster vaccinations were associated with stronger humoral and cellular immune responses.

### Antitumour activity

Among patients with evaluable RECIST assessments, 6 of 18 had stable disease (SD) and 12 had progressive disease (PD) at week 8; 2 of 6 had SD and 4 had PD at week 16; and 3 of 8 had SD and 5 had PD at week 24. The exact timing of assessments varied between patients, as detailed in the swimmer plot (Figure 5A). Across all assessment time points, the best overall response was SD in 6 of 24 patients (25.0%), PD in 17 (70.8%), and not evaluable in 1 (4.2%).

**Figure 5.**
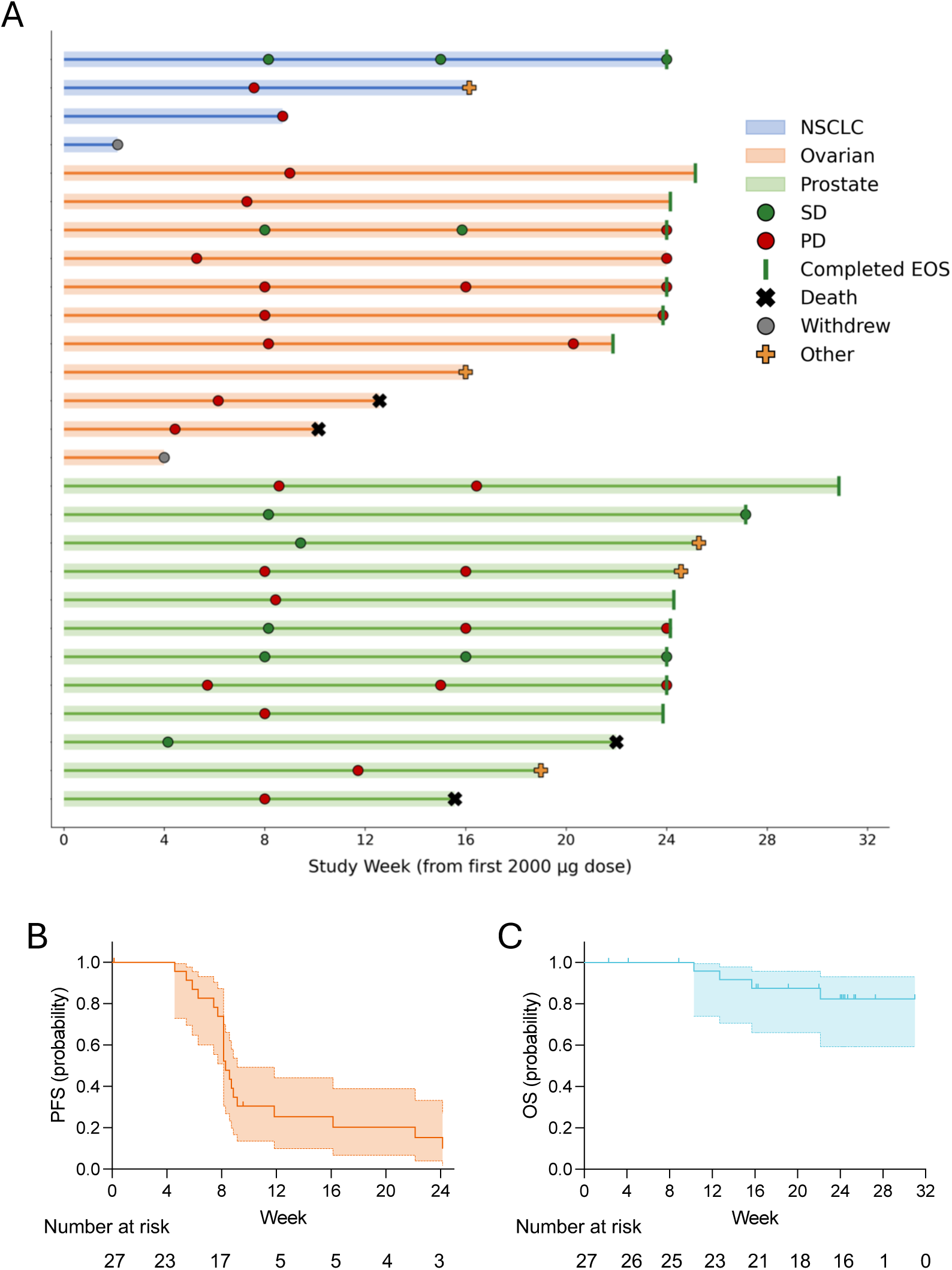
Clinical activity of OVM-200 at 2000 μg. (A) Swimmer plot showing individual patient time on study and tumour assessment outcomes in the 2000 μg population, grouped by tumour type (NSCLC, ovarian, and prostate cancer). Each horizontal bar represents an individual patient from first vaccination. Radiological assessments are shown at week 8, week 16, and week 24. (B-C) Kaplan-Meier plot of progression-free survival (PFS) (B) and overall survival (OS) (C) for 2000 μg total populations. Numbers at risk are shown below the plot. EOS, end of study; NE, not evaluable; PD: progressive disease; SD, stable disease.

At the data cut-off, 20 of 27 patients in the 2000 µg population had experienced disease progression or death, while 7 patients were censored. Median progression-free survival was 8.3 weeks. Progression-free survival rates were 73.9% at 8 weeks, 25.4% at 16 weeks, and 15.2% at 24 weeks (Figure 5B). 4 deaths had occurred by the data cut-off. Median overall survival had not been reached. Overall survival rates were 100% at 8 weeks, 87.5% at 16 weeks, and 82.4% at 24 weeks (Figure 5C).

### Clinical outcome and associated immunogenicity

Patients achieving disease stablisation developed stronger humoral immune responses than those with progressive disease. In the stable disease group, median anti-survivin antibody titres increased from 15 (IQR 10-55) at baseline to 30,720 (IQR 16,640-102,400) at the early post-vaccination assessment (week 5-8), 327,680 (IQR 87,040-491,520) at week 16, and 368,640 (IQR 35,840-655,360) at week 24 (Figure 6A). In contrast, patients with progressive disease showed lower antibody titres throughout follow-up, increasing from baseline, 60 (IQR 10-160), to 20,480 (IQR 10,240-81,920) at the early post-vaccination assessment, 163,840 (IQR 15,360-245,760) at week 16, and 163,840 (IQR 46,080-245,760) at week 24. Overall, patients with stable disease developed significantly higher anti-survivin antibody titres than those with progressive disease (p=0.0418),

**Figure 6.**
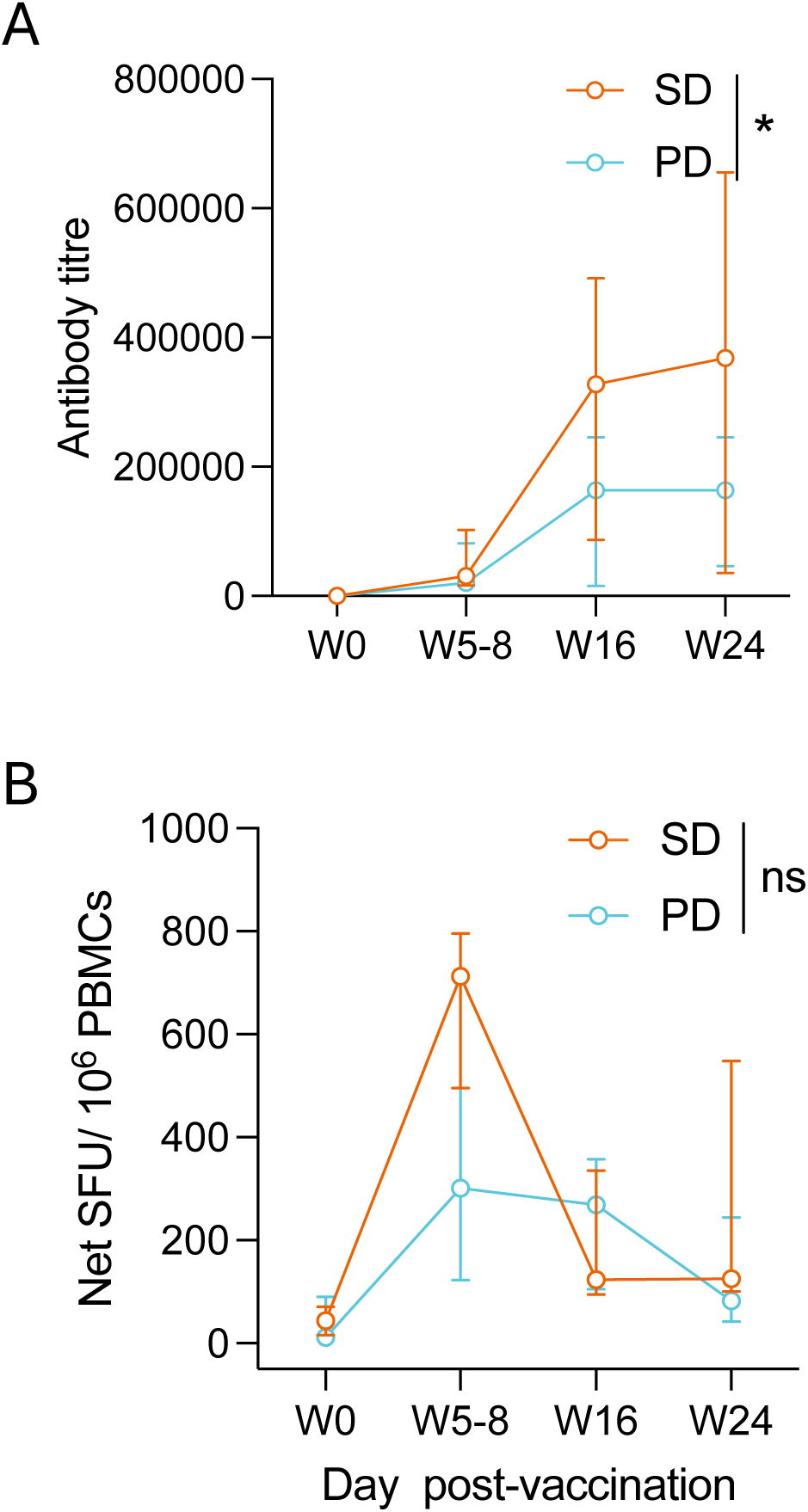
Immune responses by best overall response in patients treated with OVM-200. Comparison of survivin-specific immune responses over time between patients with stable disease and progressive disease. (A) Anti-survivin IgG titres at baseline (SD, n=6; PD, n=20), early post-vaccination (week 5-8; SD, n=6; PD, n=17), week 16 (SD, n=5; PD, n=9), and week 24 (SD, n=4; PD, n=5). (B) Interferon-γ ELISpot responses at the same timepoints (baseline: SD, n=5; PD, n=16; week 5-8: SD, n=6; PD, n=16; week 16: SD, n=5; PD, n=4; week 24: SD, n=3; PD, n=5). Data are presented as median with interquartile range (IQR). PBMCs, peripheral blood mononuclear cells; PD, progressive disease; SD, stable disease; SFU, spot-forming units.

Survivin-specific T cell responses also increased following vaccination in both groups (Figure 6B). In patients with stable disease, median ELISpot responses increased from 43.6 (IQR 15.7-71.3) SFU per 10⁶ PBMCs at baseline to 712.5 (IQR 495.3-796.1) at the early post-vaccination assessment, whereas the corresponding increase in patients with progressive disease was from 11.4 (IQR 1.0-90.3) to 301.7 (IQR 122.8-714.1) SFU per 10⁶ PBMCs. Responses remained above baseline at later time points in both groups.

### NSCLC case report

One patient with stage IV metastatic NSCLC provides an illustrative example of prolonged immune activation accompanied by durable disease stabilisation. The patient had stage IV NSCLC and had experienced disease progression following both first-line and second-line systemic therapy before enrolment into the OVM-200 trial in May 2025 (Figure 7A). The patient received 11 vaccinations, administered every 2 weeks from day 0 to day 140, and maintained stable disease at both week 8 and week 24 assessments (Figure 7B).

**Figure 7.**
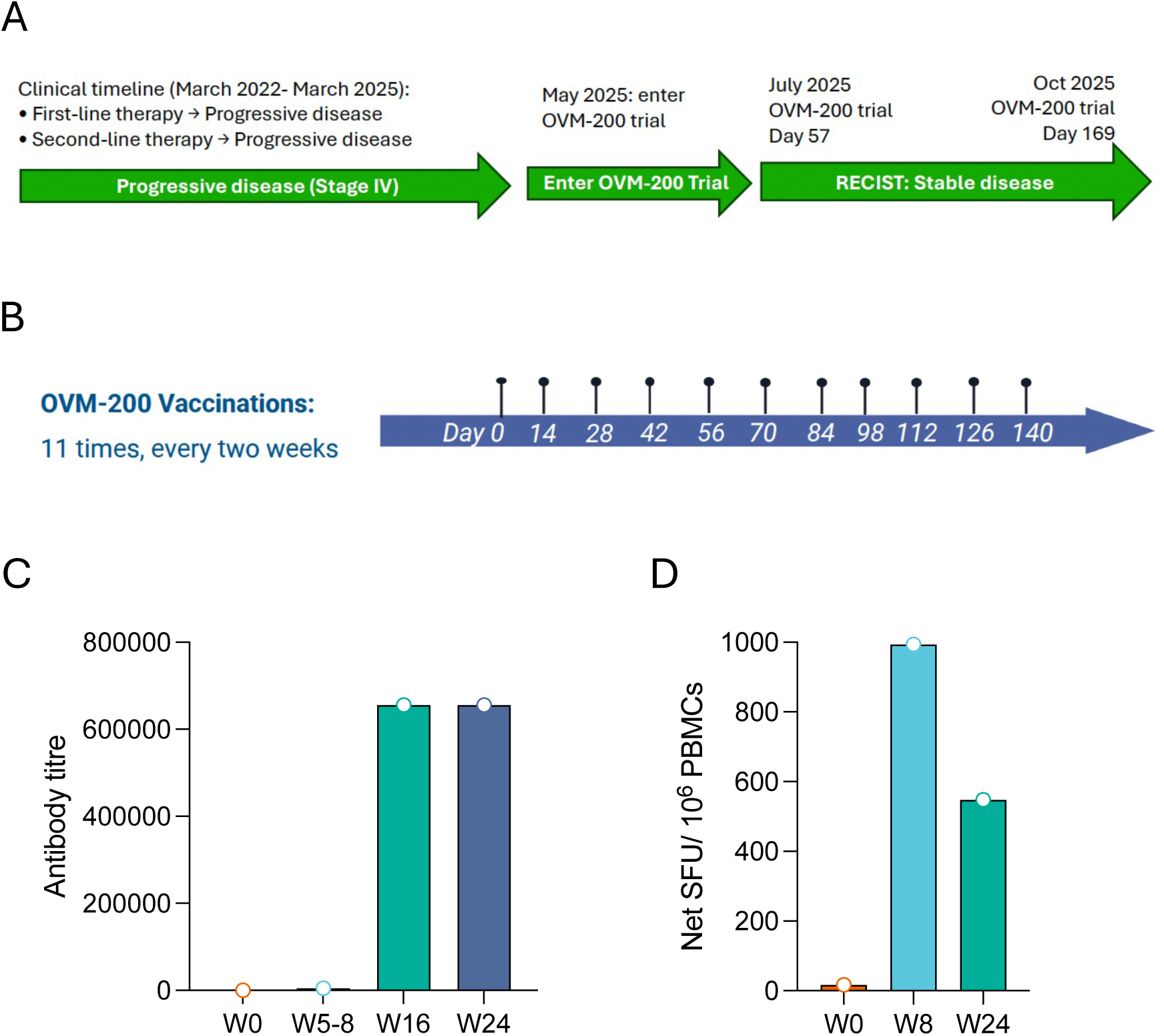
NSCLC patient case report. Clinical timeline and immune response assessment of an NSCLC patient following OVM-200 vaccination. (A) Clinical course and disease assessment timeline. (B) OVM-200 vaccination schedule. (C) Anti-survivin IgG antibody titres at the indicated timepoints. (D) Survivin-specific interferon-γ ELISpot responses at the indicated timepoints. NSCLC, non-small cell lung cancer; SFU, spot-forming units; PBMCs, peripheral blood mononuclear cells.

Anti-survivin antibody titre increased from 10 at baseline to 5120 at the early post-vaccination assessment (Week 5-8), reaching 655,360 at both Week 16 and Week 24. Survivin-specific T cell response increased from 17.4 SFU per 10⁶ PBMCs at baseline to 993.6 SFU per 10^6^ PBMCs at Week 8 and remained at 547.7 SFU per 10^6^ PBMCs at Week 24.

This case demonstrates prolonged administration of OVM-200 was associated with sustained humoral and cellular immune responses together with durable disease stabilisation in a patient whose disease had progressed despite prior standard therapy..

## Discussion

In this combined dose-expansion analysis of OVM-200 at 2000 µg, comprising of 24 patients enrolled in the phase 1b and 3 patients treated at this dose in the phase 1a, OVM-200 showed a favourable safety profile and induced survivin-specific humoral and cellular immune responses in patients with advanced NSCLC, ovarian cancer, and prostate cancer. Survivin is widely overexpressed in cancer but has limited expression in normal adult tissues^38,39^. However, as a non-mutated self-antigen, it is subject to central and peripheral immune tolerance. The induction of both antibody and T cell responses therefore provides clinical evidence that the recombinant overlapping peptide (ROP) platform can overcome functional immune tolerance and generate anti-survivin immunity in patients with advanced cancer.

Unlike conventional peptide vaccines, OVM-200 re-engineers the full-length survivin sequence into a recombinant overlapping peptide construct separated by cathepsin-sensitive linkers to facilitate antigen processing. This design offers several potential advantages^20^. First, the recombinant structure may increase immunological “foreignness” while preserving a broad repertoire of survivin epitopes. Second, because the survivin sequence is fragmented by linker sequences, the recombinant protein is unlikely to retain the native tertiary structure or anti-apoptotic biological activity of survivin, reducing concerns associated with administration of an intact oncoprotein. Third, processing of full-length antigen generates multiple peptides for presentation through both MHC class I and II pathways, supporting coordinated CD8⁺ cytotoxic and CD4⁺ helper T cell responses. A further distinguishing feature is its antigenic breadth. Earlier survivin vaccines commonly targeted selected HLA-restricted epitopes, including the HLA-A*24-restricted survivin-2B₈₀-₈₈ peptide^40^, while SurVaxM targets a defined 15-amino acid survivin region designed for presentation across multiple selected HLA backgrounds^41^. By spanning the full survivin sequence, OVM-200 may enable broader CD4⁺ and CD8⁺ T cell recognition without restriction to a predefined HLA haplotype.

OVM-200 was well tolerated, with toxicity predominantly comprising low-grade injection-site events and no dose-limiting toxicities, treatment-related serious adverse events, or treatment-related deaths. This is consistent with the phase 1a study and previous survivin vaccine trials^16,22^. Repeated administration was feasible, with selected patients receiving up to 11 vaccinations without evidence of cumulative dose-limiting toxicity, supporting the feasibility of prolonged vaccination to establish and maintain immunity.

OVM-200 induced rapid and durable anti-survivin antibody responses, with greater numbers of vaccinations associated with stronger humoral responses. Repeated antigen exposure may promote memory B cell and long-lived plasma cell responses, contributing to sustained antibody production^42,43^. In contrast, survivin-specific T cell responses were strongest at early post-vaccination timepoints and subsequently declined, potentially reflecting contraction of the effector response and transition towards memory populations^44,45^. Overall, prolonged vaccination appeared to have a greater effect on humoral than cellular immunity.

Humoral responses were also stronger in patients with stable disease, with anti-survivin antibody titres continuing to rise through week 24, whereas titres declined in patients with progressive disease. A similar association between elevated anti-survivin antibody levels and clinical outcome has been reported with SurVaxM^17^. In contrast, survivin-specific T cell responses did not significantly differ between stable and progressive disease, consistent with the SurVaxM study, in which T cell activation did not correlate with overall survival^17^. However, comparisons with our findings should be interpreted with caution, because the SurVaxM study enrolled patients with newly diagnosed glioblastoma, whereas our cohort comprised patients with extensively treated advanced solid tumours.

The ability to generate broad immunity against a shared tumour-associated antigen provides a alternative strategy to personalised neoantigen vaccination. Personalised vaccines, including mRNA-4157 (V940)^46^ and autogene cevumeran^47^, have demonstrated encouraging clinical activity but require tumour sequencing, neoantigen prediction and individualised manufacturing. In contrast, OVM-200 is an off-the-shelf vaccine that can be manufactured at scale and administered without patient-specific antigen selection. It demonstrates that appropriate engineering of tumour-associated antigens can generate potent immunity despite pre-existing immune tolerance.

Several limitations should be considered. The study had a small sample size and was non-randomised, with a relatively short follow-up period. In addition, the heavily pretreated population may have limited the ability of vaccine-induced immunity to translate into objective tumour responses. Immune responses against survivin summarised in the systematic review should also be interpreted cautiously because of differences in tumour type, prior treatment and immunological assays used to determine antibody titres in different studies.

These findings nevertheless inform future clinical development. The early peak in survivin-specific T cell responses suggests that additional immune support may be required to sustain effector function, and preclinical studies have shown that anti-4-1BB agonism can enhance the antitumour activity of survivin-based ROP vaccination^20^. Potential combinations include co-stimulatory agonists, immune checkpoint blockade, T cell-engaging therapies and cytokine-based approaches. In parallel, therapeutic cancer vaccine development has increasingly shifted towards earlier disease settings, particularly following surgery or other curative-intent treatment^46,47^. SurVaxM has similarly been evaluated into newly diagnosed glioblastoma following surgery and chemoradiotherapy. In these settings, lower tumour burden and reduced tumour-associated immune suppression may favour vaccine-induced elimination of residual tumour cells. These observations provide a strong scientific rationale for evaluating OVM-200 in earlier disease settings, such as the adjuvant and neoadjuvant settings, where induction of durable survivin-specific immunity may be better positioned to eliminate residual tumour cells and reduce the risk of recurrence.

In summary, OVM-200 at 2000 μg showed a favourable safety profile and induced survivin-specific humoral and cellular immune responses in patients with advanced solid tumours who had exhausted available standard therapies. Together, these findings provide clinical proof-of-mechanism evidence for the ROP approach and support continued development of OVM-200.

## Methods

### Study design and participants

OVM-200-100 was a first-in-human, multicentre, open-label, phase 1 study comprising a phase 1a dose-escalation followed by a phase 1b dose-expansion. This study focuses on patients who received OVM-200 at 2000 μg, comprising those treated at 2000 μg during phase 1a (n=3) and the full phase 1b expansion cohort (n=24), for a combined 2000 μg analysis population of 27 patients. The dose was selected on the basis of the phase 1a component of the study. The phase 1b was designed to further assess the safety, tolerability, immunogenicity, and preliminary antitumour activity of OVM-200 in patients with advanced NSCLC, ovarian cancer, and prostate cancer. The study was conducted at four sites in the UK. Trial registration identifiers were ClinicalTrials.gov NCT05104515 and EudraCT 2021-001545-12. Recruitment and follow-up took place between 15 Oct 2021 and 3 Dec 2025.

Eligible patients were aged 18-75 years and had histologically confirmed metastatic or locally advanced inoperable NSCLC, ovarian cancer, or prostate cancer, with at least one previous line of approved anticancer therapy. Patients were required either to have exhausted recognised treatment options, to be in a planned treatment-free interval after a defined course of therapy, or, in the case of prostate cancer, to be clinically stable on anti-hormonal treatment. Active anti-cancer therapy at enrolment was not permitted, apart from supportive treatment and androgen-deprivation therapy for prostate cancer. For NSCLC, at least one measurable lesion suitable for repeated radiological assessment was required. Additional eligibility criteria included life expectancy of at least 3 months and adequate haematological, renal, and hepatic function.

Key exclusion criteria included clinically significant immunodeficiency; systemic corticosteroids exceeding prednisolone 10 mg/day equivalent or other immunosuppressive medication within 14 days before first dose; active or suspected autoimmune disease requiring systemic therapy; prior anticancer vaccine therapy, immune checkpoint antibody or T cell co-stimulation therapy within 28 days before first dose; investigational treatment within 28 days or 6 half-lives, whichever was longer; major surgery, chemotherapy, or radiotherapy within 28 days before first dose; active infection or viral disease; any vaccine within 28 days before first dose; symptomatic brain metastases or leptomeningeal disease; uncontrolled medical disorders; or known hypersensitivity to any component of OVM-200 or the adjuvant. Survivin expression was not an entry criterion and was assessed only in a subset of patients as an exploratory biomarker.

The study protocol was approved by the South Central-Oxford A Research Ethics Committee (22/SC/0342). The study was conducted in accordance with the Declaration of Helsinki, Good Clinical Practice guidelines, and UK clinical trial regulations. All participants provided written informed consent.

### Procedures

OVM-200 is a recombinant overlapping peptide vaccine spanning the full-length survivin sequence and formulated with Montanide ISA 51 VG. Vaccine material was manufactured under Good Manufacturing Practice conditions by Eurofins CDMO (Ghent, Belgium). The final product was supplied as a 2 mg/mL solution and emulsified 1:1 with Montanide before subcutaneous administration.

All patients received OVM-200 2000 μg subcutaneously every 2 weeks for an initial series of three vaccinations. Patients were observed for at least 4 h after each administration. Patients considered by the investigator to be deriving benefit could continue treatment for up to eight additional immunisations, to a maximum of 11 doses. Baseline assessments were performed before the first vaccination, with on-treatment evaluation through the end-of-treatment visit at week 5 (day 36), and follow-up assessments at week 8 (day 57), week 16 (day 113), and end of study at week 24 (day 169).

Blood for immunogenicity analyses was collected at baseline (day 1), week 5 (day 36), week 8 (day 57), week 16 (day 113), and week 24 (day 169). Serum anti-survivin IgG responses were measured by ELISA, with titres reported in two-fold dilutions starting at 1:20. Titres below 1:20 were assigned a value of 10 (half the lower limit of quantification) for geometric mean calculations. Survivin-specific cellular responses were assessed by interferon-γ ELISpot using peripheral blood mononuclear cells isolated from whole blood within 24 h of collection.

Tumour assessments were performed during study follow-up according to RECIST version 1.1, with week 8 and later assessments contributing to best overall response. Safety assessments included adverse-event monitoring, physical examination, ECOG performance status, vital signs, laboratory testing, urinalysis, and electrocardiograms. Adverse events were graded according to CTCAE version 5.0.

### Outcomes

The primary endpoint was the safety and tolerability of OVM-200 at 2000 μg, assessed by the occurrence, severity, and relationship of adverse events, including dose-limiting toxicities, serious adverse events, and adverse events leading to treatment discontinuation.

Secondary endpoints were survivin-specific immunogenicity, assessed by anti-survivin IgG ELISA and interferon-γ ELISpot; and the association between the number of vaccinations received and survivin-specific immunogenicity.

Exploratory endpoints included associations between immunogenicity and clinical outcome; preliminary antitumour activity, assessed by best overall response according to RECIST version 1.1; progression-free survival; associations between survivin expression in tumour and survivin-specific immunogenicity; and durability of vaccine-induced immune responses.

### Statistical analysis

The safety analysis comprised all patients who received at least one dose of OVM-200 (n=27). Immunogenicity and antitumour activity were assessed in evaluable patients with available data for the relevant endpoint. Data are presented as median (IQR) for continuous variables and n (%) for categorical variables. The IQR was defined by the 25th and 75th percentiles using the Type 6 quantile method. Where only two observations were available, plotted error bars represent the full range (minimum and maximum). The objective response rate is presented with 95% confidence intervals. Comparisons between two groups were performed using t-tests where appropriate. Immunogenicity responses over time were compared between patients with stable disease and those with progressive disease using two-way ANOVA with multiple-comparisons correction. Statistical significance is indicated as *p < 0.05, **p < 0.01, ***p < 0.001, and ****p < 0.0001. All statistical tests were two-sided. All analyses were performed using GraphPad Prism version 9.

Seroprevalence data pooled from published studies are presented both as the median of study-level proportions with range, and as a patient-weighted pooled proportion. The number of contributing studies and total patients are stated.

## Data Availability

Requests for data sharing will be reviewed on a case-by-case basis and subject to data sharing agreements.

## Contributions

PKTW and SJ wrote the first draft of the manuscript. PKTW, TM, TZ, and SJ contributed to the data curation. PKTW and TZ undertook the formal data analysis and visualisation. PKTW, TM, WF, and SJ had full access to all the data in the study. TM, WF, and SJ verified the data in the study and were responsible in the supervision of the trial. MF, FC, MT, FT, and AW were responsible for the recruitment and management of the patients in their respective trial site. All authors contributed to data interpretation, drafting and review of the final manuscript, and had final responsibility for the decision to submit for publication.

## Data sharing statement

Individual patient data from this trial will not be published in the public domain. Requests for data sharing will be reviewed on a case-by-case basis and subject to data sharing agreements. The trial protocol and statistical analysis plan are available in Supplementary Information.

## Acknowledgements

We thank all patients who participated in this clinical trial. We also thank all staff members at the trial sites for their invaluable contributions. We acknowledge Fortrea as the contract research organisation (CRO) for their contributions to trial coordination, data management, and immunological testing. This work was supported by Oxford Vacmedix UK Ltd.

---

**Supplementary Figure 1.**
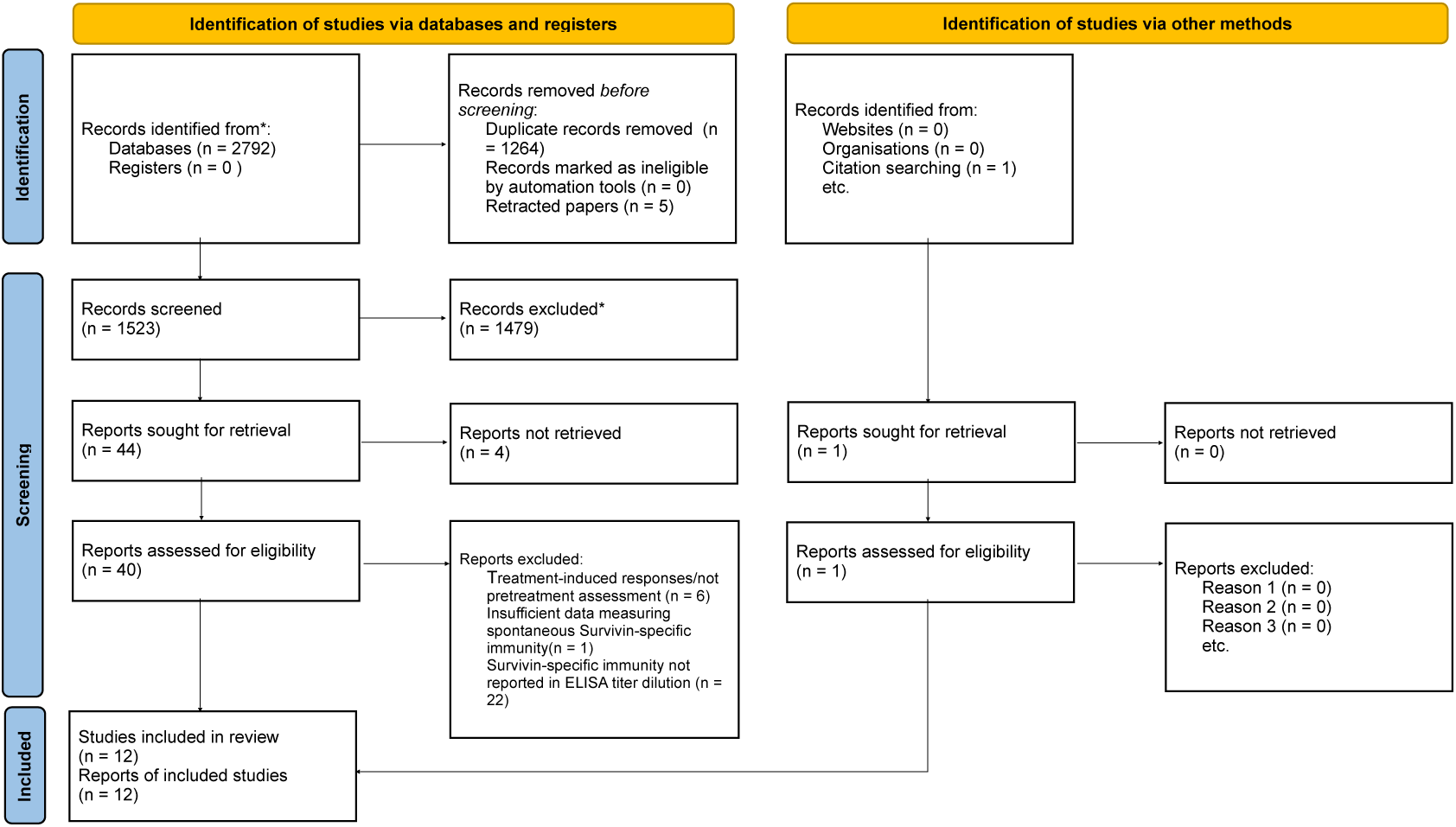
PRISMA flow diagram of the systematic literature search. PRISMA 2020 flow diagram showing the identification, screening, eligibility assessment and inclusion of studies evaluating spontaneous survivin-specific immunity. Records were identified through database searches and citation searching, with 12 studies included in the final analysis.

**Supplementary Table 1.** Treatment-emergent adverse events in patients. Treatment-emergent adverse events (TEAEs) reported in patients who received OVM-200 2000 µg (n=27), summarised by system organ class and preferred term. Data are n (%). Preferred terms reported in ≥2 patients are shown. Grade 3 events are included irrespective of frequency.

| **Treatment-Emergent Adverse Events (TEAEs)** | **Grade 1–2** | **Grade 3** |
|---|---|---|
| **Summary** |  |  |
| Any TEAE† | 21 (78%) | 6 (22%) |
| OVM-200-related TEAE | 27 (100%) | 0 |
| Leading to discontinuation of OVM-200 | 1 (4%) | 0 |
| No dose-limiting toxicities | 0 | 0 |
| **General disorders & administration site conditions** |  |  |
| Injection site reaction | 17 (63%) | 0 |
| Injection site erythema | 6 (22%) | 0 |
| Fatigue | 6 (22%) | 0 |
| Injection site pruritus | 3 (11%) | 0 |
| Pyrexia | 3 (11%) | 0 |
| Injection site pain | 2 (7%) | 0 |
| Peripheral swelling | 2 (7%) | 0 |
| **Gastrointestinal disorders** |  |  |
| Vomiting | 4 (15%) | 1 (4%) |
| Diarrhoea | 4 (15%) | 0 |
| Nausea | 2 (7%) | 1 (4%) |
| Abdominal pain | 2 (7%) | 1 (4%) |
| Constipation | 2 (7%) | 0 |
| Ascites | 0 | 1 (4%) |
| **Musculoskeletal & connective tissue disorders** |  |  |
| Arthralgia | 4 (15%) | 0 |
| Pain in extremity | 3 (11%) | 0 |
| Back pain | 3 (11%) | 0 |
| Myalgia | 3 (11%) | 0 |
| **Respiratory, thoracic & mediastinal disorders** |  |  |
| Dyspnoea | 4 (15%) | 0 |
| Cough | 2 (7%) | 0 |
| **Metabolism & nutrition disorders** |  |  |
| Hypoalbuminaemia | 1 (4%) | 1 (4%) |
| Decreased appetite | 2 (7%) | 0 |
| Iron deficiency | 2 (7%) | 0 |
| Dehydration | 0 | 1 (4%) |
| **Investigations** |  |  |
| Blood magnesium decreased | 2 (7%) | 0 |
| Gamma-glutamyltransferase increased | 0 | 2 (7%) |
| **Blood & lymphatic system disorders** |  |  |
| Anaemia | 3 (11%) | 1 (4%) |
| **Nervous system disorders** |  |  |
| Headache | 4 (15%) | 0 |
| **Reproductive system & breast disorders** |  |  |
| Pelvic pain | 1 (4%) | 1 (4%) |
| **Infections & infestations** |  |  |
| Vascular device infection | 0 | 1 (4%) |
| Pneumonia | 0 | 1 (4%) |
| Peritonitis | 0 | 1 (4%) |
| **Vascular disorders** |  |  |
| Hypotension | 0 | 1 (4%) |

†5 patients (19%) experienced serious TEAEs, comprising 9 events in total. None were considered related to OVM-200.

**Supplementary Table 2.** Reported spontaneous anti-survivin antibody seroprevalence in patients with cancer not receiving survivin-directed immunotherapy. The 12 studies are grouped by serum dilution, with the number of patients, number of survivin-antibody-positive patients, and percentage positivity reported for each study.

| **Study** | **Dilution** | **N** | **Positive** | **% Positive** | **PMID/ Note** |
|---|---|---|---|---|---|
| Zörnig 2014 | 1:50 | 365 | 41 | 11.2% | *PMID: 24839182* |
| **Subtotal, 1:50** |  | **365** | **41** | **11.2%** | **1 study** |
| Yagihashi 2004 | 1:100 | 31 | 18 | 58.1% | *PMID: 15829321* |
| Li 2024 | 1:100 | 368 | 117 | 31.8% | *PMID: 38832035* |
| Wang 2012 | 1:100 | 135 | 42 | 31.1% | *PMID: 23317217* |
| Rohayem 2000 | 1:100 | 100 | 15 | 15.0% | *PMID: 10766164* |
| Meistere 2017 | 1:100 | 18 | 3 | 16.7% | *PMID: 28768706* |
| Karanikas 2009 | 1:100 | 150 | 9 | 6.0% | *PMID: 18789878* |
| Söling 2006* | 1:100 | 77 | 8 | 10.4% | *PMID: 17143520* |
| **Subtotal, 1:100** |  | **879** | **212** | **24.1%** | **7 studies; median 16.7%** |
| Zhang 2006 | 1:200 | 142 | 16 | 11.3% | *PMID: 17289549* |
| Dai 2014 | 1:200 | 174 | 35 | 20.1% | *PMID: 24860838* |
| Wang 2014 | 1:200 | 304 | 48 | 15.8% | *PMID: 24822225* |
| Megliorino 2005 | 1:200 | 1137 | 96 | 8.4% | *PMID: 15896923* |
| **Subtotal, 1:200** |  | **1757** | **195** | **11.1%** | **4 studies; median 13.5%** |
| **Total** |  | **3001** | **448** | **14.9%** | **12 studies; median 15.4%** |

\* 1/77 patient is positive for survivin antibody at 1:1000 dilution

## References

1 Saxena, M., van der Burg, S. H., Melief, C. J. M. & Bhardwaj, N. Therapeutic cancer vaccines. Nat Rev Cancer 21, 360–378 (2021). 10.1038/s41568-021-00346-0

2 Braun, D. A. et al. A neoantigen vaccine generates antitumour immunity in renal cell carcinoma. Nature 639, 474–482 (2025). 10.1038/s41586-024-08507-5

3 Rojas, L. A. et al. Personalized RNA neoantigen vaccines stimulate T cells in pancreatic cancer. Nature 618, 144–150 (2023). 10.1038/s41586-023-06063-y

4 Yarchoan, M. et al. Personalized neoantigen vaccine and pembrolizumab in advanced hepatocellular carcinoma: a phase 1/2 trial. Nature Medicine 30, 1044–1053 (2024). 10.1038/s41591-024-02894-y

5 Sahin, U. et al. Individualized mRNA vaccines evoke durable T cell immunity in adjuvant TNBC. Nature 651, 1088–1096 (2026). 10.1038/s41586-025-10004-2

6 Fan, T. et al. Therapeutic cancer vaccines: advancements, challenges and prospects. Signal Transduction and Targeted Therapy 8, 450 (2023). 10.1038/s41392-023-01674-3

7 Gulley, J. L. et al. Phase III Trial of PROSTVAC in Asymptomatic or Minimally Symptomatic Metastatic Castration-Resistant Prostate Cancer. J Clin Oncol 37, 1051–1061 (2019). 10.1200/jco.18.02031

8 Middleton, G. et al. Gemcitabine and capecitabine with or without telomerase peptide vaccine GV1001 in patients with locally advanced or metastatic pancreatic cancer (TeloVac): an open-label, randomised, phase 3 trial. Lancet Oncol 15, 829–840 (2014). 10.1016/s1470-2045(14)70236-0

9 Chen, X., Duan, N., Zhang, C. & Zhang, W. Survivin and Tumorigenesis: Molecular Mechanisms and Therapeutic Strategies. J Cancer 7, 314–323 (2016). 10.7150/jca.13332

10 Asanuma, H. et al. Survivin expression is regulated by coexpression of human epidermal growth factor receptor 2 and epidermal growth factor receptor via phosphatidylinositol 3-kinase/AKT signaling pathway in breast cancer cells. Cancer Res 65, 11018–11025 (2005). 10.1158/0008-5472.Can-05-0491

11 Altieri, D. C. Survivin, cancer networks and pathway-directed drug discovery. Nature Reviews Cancer 8, 61–70 (2008). 10.1038/nrc2293

12 Mita, A. C., Mita, M. M., Nawrocki, S. T. & Giles, F. J. Survivin: Key Regulator of Mitosis and Apoptosis and Novel Target for Cancer Therapeutics. Clinical Cancer Research 14, 5000–5005 (2008). 10.1158/1078-0432.Ccr-08-0746

13 Fukuda, S. & Pelus, L. M. Survivin, a cancer target with an emerging role in normal adult tissues. Molecular Cancer Therapeutics 5, 1087–1098 (2006). 10.1158/1535-7163.Mct-05-0375

14 Goodnow, C. C., Sprent, J., de St Groth, B. F. & Vinuesa, C. G. Cellular and genetic mechanisms of self tolerance and autoimmunity. Nature 435, 590–597 (2005). 10.1038/nature03724

15 Lennerz, V. et al. Immunologic response to the survivin-derived multi-epitope vaccine EMD640744 in patients with advanced solid tumors. Cancer Immunol Immunother 63, 381–394 (2014). 10.1007/s00262-013-1516-5

16 Fenstermaker, R. et al. IT-09PHASE I STUDY OF SAFETY, TOLERABILITY AND IMMUNOLOGIC EFFECTS OF A SURVIVIN PEPTIDE MIMIC VACCINE (SurVaxM) IN PATIENTS WITH RECURRENT MALIGNANT GLIOMA. Neuro-Oncology 16, v111-v111 (2014). 10.1093/neuonc/nou258.7

17 Ahluwalia, M. S. et al. Phase IIa Study of SurVaxM Plus Adjuvant Temozolomide for Newly Diagnosed Glioblastoma. Journal of Clinical Oncology 41, 1453–1465 (2023). 10.1200/jco.22.00996

18 Shen, H. et al. Identification of a novel HLA-A2-restricted mutated Survivin epitope and induction of specific anti-HCC CTLs that could effectively cross-recognize wild-type Survivin antigen. Cancer Immunol Immunother 62, 393–403 (2013). 10.1007/s00262-012-1323-4

19 Ogasawara, M. et al. Induction of Survivin-Specific, HLA-A24 Restricted Cytotoxic T Lymphocytes Using an Immunodominant Peptide; In-Vitro Feasibility Study and a Clinical Trial of Survivin Peptide Vaccination. Blood 108, 2709 (2006). 10.1182/blood.V108.11.2709.2709

20 Zhang, Y. et al. Survivin-Based Recombinant Overlapping Peptides Induce T Lymphocyte Cytotoxicity and Prolong the Survival in In Vivo Melanoma Model. Advanced Therapeutics 6, 2300253 (2023). 10.1002/adtp.202300253

21 Cai, L. et al. Protective cellular immunity generated by cross-presenting recombinant overlapping peptide proteins. Oncotarget 8, 76516–76524 (2017). 10.18632/oncotarget.20407

22 Wijaya, W. et al. Survivin recombinant overlapping peptide (ROP) vaccine in advanced solid tumours: a first-in-human, multicentre, open-label, phase 1a dose-escalation study. eClinicalMedicine 91 (2026). 10.1016/j.eclinm.2025.103717

23 Page, M. J. et al. The PRISMA 2020 statement: an updated guideline for reporting systematic reviews. BMJ 372, n71 (2021). 10.1136/bmj.n71

24 Zörnig, I. et al. Prognostic significance of spontaneous antibody responses against tumor-associated antigens in malignant melanoma patients. Int J Cancer 136, 138–151 (2015). 10.1002/ijc.28980

25 Yagihashi, A. et al. Detection of autoantibodies to livin and survivin in Sera from lung cancer patients. Lung Cancer 48, 217–221 (2005). 10.1016/j.lungcan.2004.11.002

26 Li, Q. et al. Anti-BIRC5 autoantibody serves as a valuable biomarker for diagnosing AFP-negative hepatocellular carcinoma. PeerJ 12, e17494 (2024). 10.7717/peerj.17494

27 Wang, Y. Q. et al. Correlation between auto-antibodies to survivin and MUC1 variable number tandem repeats in colorectal cancer. Asian Pac J Cancer Prev 13, 5557–5562 (2012). 10.7314/apjcp.2012.13.11.5557

28 Rohayem, J. et al. Antibody response to the tumor-associated inhibitor of apoptosis protein survivin in cancer patients. Cancer Res 60, 1815–1817 (2000).

29 Meistere, I. et al. The Prevalence of Cancer-Associated Autoantibodies in Patients with Gastric Cancer and Progressive Grades of Premalignant Lesions. Cancer Epidemiol Biomarkers Prev 26, 1564–1574 (2017). 10.1158/1055-9965.Epi-17-0238

30 Söling, A. et al. Autoantibodies to the inhibitor of apoptosis protein survivin in patients with brain tumors. Int J Oncol 30, 123–128 (2007).

31 Zhang, J. Y. Mini-array of multiple tumor-associated antigens to enhance autoantibody detection for immunodiagnosis of hepatocellular carcinoma. Autoimmun Rev 6, 143–148 (2007). 10.1016/j.autrev.2006.09.009

32 Dai, L. et al. Preferential autoimmune response in prostate cancer to cyclin B1 in a panel of tumor-associated antigens. J Immunol Res 2014, 827827 (2014). 10.1155/2014/827827

33 Wang, P. et al. Evaluation of diagnostic value in using a panel of multiple tumor-associated antigens for immunodiagnosis of cancer. J Immunol Res 2014, 512540 (2014). 10.1155/2014/512540

34 Megliorino, R. et al. Autoimmune response to anti-apoptotic protein survivin and its association with antibodies to p53 and c-myc in cancer detection. Cancer Detect Prev 29, 241–248 (2005). 10.1016/j.cdp.2005.03.002

35 Karanikas, V. et al. Baseline levels of CD8+ T cells against survivin and survivin-2B in the blood of lung cancer patients and cancer-free individuals. Clin Immunol 129, 230–240 (2008). 10.1016/j.clim.2008.07.024

36 Becker, J. C. et al. Survivin-specific T-cell reactivity correlates with tumor response and patient survival: a phase-II peptide vaccination trial in metastatic melanoma. Cancer Immunol Immunother 61, 2091–2103 (2012). 10.1007/s00262-012-1266-9

37 Coughlin, C. M. et al. Immunosurveillance and Survivin-Specific T-Cell Immunity in Children With High-Risk Neuroblastoma. Journal of Clinical Oncology 24, 5725–5734 10.1200/JCO.2005.05.3314

38 Kennedy, S. M. et al. Prognostic importance of survivin in breast cancer. British Journal of Cancer 88, 1077–1083 (2003). 10.1038/sj.bjc.6600776

39 Sarela, A. I. et al. Expression of survivin, a novel inhibitor of apoptosis and cell cycle regulatory protein, in pancreatic adenocarcinoma. British Journal of Cancer 86, 886–892 (2002). 10.1038/sj.bjc.6600133

40 Tsuruma, T. et al. Phase I clinical study of anti-apoptosis protein, survivin-derived peptide vaccine therapy for patients with advanced or recurrent colorectal cancer. Journal of Translational Medicine 2, 19 (2004). 10.1186/1479-5876-2-19

41 Fenstermaker, R. A. et al. Clinical study of a survivin long peptide vaccine (SurVaxM) in patients with recurrent malignant glioma. Cancer Immunol Immunother 65, 1339–1352 (2016). 10.1007/s00262-016-1890-x

42 Cohen, K. W., et al. Longitudinal analysis shows durable and broad immune memory after SARS-CoV-2 infection with persisting antibody responses and memory B and T&#xa0;cells. Cell Reports Medicine 2 (2021). 10.1016/j.xcrm.2021.100354

43 Van Beek, M., Nussenzweig, M. C. & Chakraborty, A. K. Two complementary features of humoral immune memory confer protection against the same or variant antigens. Proceedings of the National Academy of Sciences 119, e2205598119 (2022). doi:10.1073/pnas.2205598119

44 Kaech, S. M., Wherry, E. J. & Ahmed, R. Effector and memory T-cell differentiation: implications for vaccine development. Nature Reviews Immunology 2, 251–262 (2002). 10.1038/nri778

45 Benoit, J. M. et al. No evidence of immune exhaustion after repeated SARS-CoV-2 vaccination in vulnerable and healthy populations. Nature Communications 16, 5219 (2025). 10.1038/s41467-025-60216-3

46 Weber, J. S. et al. Individualised neoantigen therapy mRNA-4157 (V940) plus pembrolizumab versus pembrolizumab monotherapy in resected melanoma (KEYNOTE-942): a randomised, phase 2b study. Lancet 403, 632–644 (2024). 10.1016/s0140-6736(23)02268-7

47 Sethna, Z. et al. RNA neoantigen vaccines prime long-lived CD8+ T cells in pancreatic cancer. Nature 639, 1042–1051 (2025). 10.1038/s41586-024-08508-4

